# *De novo* variants of *NALCN* differentially impact both the phenotypic spectrum of patients and the biophysical properties of the NALCN current

**DOI:** 10.1101/2025.06.20.25329825

**Authors:** Nawale Hadouiri, Lidiane Pereira Garcia, Romain Baudat, Paloma Parra-Díaz, Antonio Gil-Nagel Rein, Isabel Del Pino, Sandra Whalen, Sarah Grotto, Theresa Brunet, Melanie Brugger, Dana Marafi, Katharina Vill, Damien Lederer, Deniz Karadurmus, Julie Desir, Marie Cécile Nassogne, Maud Favier, Siddharth Srivastava, Elise Brischoux Boucher, Jonathan Levy, Dana Young, Gabriella Horvath, Isabelle Marey, Klaus Dieterich, Chiara Fiorillo, Heike Weigand, Nora Hannane, Amelle Shillington, Lila Stange, Aditi Dagli, Emanuela Argilli, Carolyn Le, Elliott H. Sherr, Bo Hoon Lee, Ryan W Gates, Isabelle Maystadt, Marie Deprez, Gaetan Lesca, Gilles Rode, Valentin Ruault, Luca Soliani, Evamaria Lanzarini, Alison J. Eaton, François D. Morneau-Jacob, Gillian Prinzing, Annapurna Poduri, Stephanie Sacharow, Stephen R. Chorney, Elisa Rahikkala, Vasileiou Georgia, André Reis, Melissa Pauly, Ulrike Huffmeier, Cornelia Kraus, Christopher P. Barnett, Hamish S Scott, Daniel Calame, Jeremy A. Tanner, Egidio Spinelli, Frédéric Tran Mau-Them, Mathieu Gueugnon, François Lebeaupin, Antonio Vitobello, Laurence Faivre, Philippe Lory, Christel Thauvin-Robinet, Arnaud Monteil

## Abstract

The Na^+^ leak channel NALCN regulates the resting membrane potential and consequently cell excitability of several cell types, including neurons. Studies of animal models demonstrated that NALCN is involved in fundamental physiological functions such as respiratory rhythm, circadian rhythm, sleep, locomotor behavior and pain perception. Pathogenic variants of *NALCN* have been associated with ultra-rare developmental disorders characterized by a wide range of symptoms with variable severity. We and others previously showed that pathogenic variants of *NALCN* can be categorized in 2 groups. The first group corresponds to inherited biallelic loss-of-function variants with patients suffering from the IHPRF1 syndrome (OMIM #615419). The second one corresponds to *de novo* gain-of-function variants that cause the CLIFAHDD syndrome (OMIM #616266). In this study, we provide a standardized phenotypic description of a large group of 35 individuals with *de novo* pathogenic variants of *NALCN*. In addition, we performed functional studies of several of these variants using the patch clamp technique in a recombinant system. We highlight a large heterogeneity in terms of both expressed symptoms and their severity. By contrast with previous reports only showing a pure gain-of-function effect of *de novo* pathogenic variants, we found that *de novo* variants of *NALCN* differentially impact the biophysical properties of the NALCN current and likely influence cell excitability. To conclude, *de novo* variants of *NALCN* differentially impact the biophysical properties of the NALCN current. We hypothesize that this may at least partly explain the phenotypic diversity observed in patients.

## INTRODUCTION

The *NALCN* gene extends on a large genomic region around 363 kb located in 13q33.1 in humans and contains at least 44 exons. It encodes for a Na^+^ leak channel that belongs to the 4-domain ion channel family (1,2). *NALCN* is mainly expressed in the central nervous system (1,3), heart, endocrine glands and myometrial cells during labor (4,5). NALCN contributes to the regulation of the resting membrane potential (RMP) influencing cell excitability of several cell types such as neurons, endocrine cells and interstitial cells of Cajal (*reviewed in* (6–8)). Several studies using animal models reported that *NALCN* is involved in several fundamental physiological processes (6,8) such as respiratory rhythm (9), circadian rhythm (10), locomotor activity (11), pain sensitivity (12), sleep (13), gastrointestinal motility (14), systemic osmoregulation (15) and parturition (16).

The Infantile Hypotonia with Psychomotor Retardation and characteristic Facies type 1 (IHPRF1, OMIM #615419) and the Congenital contractures of the LImbs and FAce, hypotonia, and Developmental Delay (CLIFAHDD, OMIM #616266) syndromes are severe and ultra-rare genetic disorders with an onset in infancy. The IHPRF1 syndrome is caused by inherited biallelic *NALCN* variants (17–22) while the CLIFAHDD is caused by *de novo* heterozygous pathogenic variants of *NALCN*. These syndromes are characterized by a wide range of symptoms with variable severity (*reviewed in* (6)). These include facial dysmorphism, developmental delay, intellectual disability, seizures, episodic or cerebellar ataxia and severe respiratory disturbances, digestive impairment and in most severe cases premature death (17,21–32). More than 40 patients have been described so far for each syndrome (6). Unfortunately, there are no therapeutic options clearly identified to date to treat these patients (6). While most of the biallelic variants of *NALCN* described so far in patients with the IHPRF1 syndrome are predicted to be loss-of-function, by resulting either in truncated and non-functional forms of NALCN or in a mRNA decay process, *de novo* variants of *NALCN* found in patients with the CLIFAHDD syndrome were functionally described as leading to a gain-of-function (33,34). In this study, we report the clinical features of a cohort of 35 cases with *de novo NALCN* variants to better evaluate the broad phenotypic spectrum of NALCN patients. We also performed *in vitro* electrophysiological recordings in recombinant system to evaluate the functional impacts of *de novo* variants on the NALCN current. In this work, we tested the hypothesis that there may be different biophysical consequences of *de novo NALCN* variants on the NALCN current, leading to phenotypic variability.

## MATERIAL AND METHODS

### Patients

We firstly identified by exome sequencing the *de novo* pathogenic NM_052867.2:c.4004G>A *NALCN* missense variant (p.S1335N) in an individual in early adolescence with mild dysmorphic features, global developmental disorders, episodic ataxia, ataxic gait, cerebellar atrophy but without any contractures of the limbs. Through national and international data sharing, we recruited 34 additional individuals with a *de novo NALCN* variant through a collaboration with patients advocacy groups (*i.e.,* Libellas foundation, https://fundacionlibellas.org/, and Channeling Hope foundation, https://www.channelinghope.org/) as well as from an international collaboration with GeneMatcher (https://genematcher.org/) (35).

### Clinical features and molecular assessment of the patients

Each clinician (geneticist, neurologist, pediatrician or physiatrist) in each collaborating center used a standardized clinical sheet to provide phenotypic details for individuals from their center with *de novo NALCN* variations after an exhaustive review of the literature (*see* **Supplementary Table 1**). The type of contractures was specified, *i.e.*, if (i) contractures were isolated or multiple (as arthrogryposis multiplex congenita, distal arthrogryposis involving joint contractures that restricted movement in the hands and feet), and if (ii) contractures affected small and distal joints (fingers, camptodactyly, ulnar deviation, adducted thumbs, clubfoot) or large joints (hips, elbows, knees). Written informed consent was obtained from all participants in accordance with the local ethics committee to publish their phenotypic details; and for some participants to publish their clinical images and key brain MRI. The individuals were diagnosed by singleton exome sequencing (ES) with familial segregation analysis by Sanger sequencing, excepted the individuals 1, 2, 3, 4, 20 and 31, which were diagnosed using trio ES. Variant interpretation and classification was done adhering to American College of Medical Genetics and Genomics/Association for Molecular Pathology guidelines (36).

### DNA constructs

Human NALCN fused to eGFP at its carboxy-terminus, UNC-79, UNC-80 and FAM155A cDNAs cloned in the mammalian expression plasmid pcDNA3.1(+) (Thermo Fisher Scientific #V79020) were kindly provided by Dr H.C. Chua and Pr S.A. Pless (37). Pathogenic variants of NALCN were generated by gene synthesis (Genscript). Integrity of cDNAs was carefully checked by DNA sequencing following amplification (Eurofins Genomics).

### Cell culture

HEK-293T cells, also referred to as tsA201 cells, were obtained from the European Collection of Authenticated Cell Cultures (ECACC #96121229). The identity of HEK-293T has been confirmed by STR profiling and the cells have been eradicated from mycoplasma at ECACC. We routinely tested the cells for the absence of any mycoplasma contamination. Cells were cultivated at 37°C in DMEM supplemented with GlutaMax (Thermo Fisher Scientific #31966047), 10% fetal bovine serum (Thermo Fisher Scientific #10270106) and 1% penicillin/streptomycin (Thermo Fisher Scientific #15140122).

### Transfections

Transfections were performed in 35 mm petri dishes using the jetPEI reagent (Polyplus #101000053) with 2 µg of a DNA mix containing pcDNA3-NALCN-GFP (wild-type (WT) or mutant), pcDNA3-UNC-79, pcDNA3-UNC-80 and pcDNA3-FAM155A in a 1:1:1:1 molar ratio accordingly to the manufacturer protocol. pcDNA3-NALCN-GFP was replaced by a home-made pcDNA3-GFP plasmid in control conditions.

### Electrophysiological recordings

Transfected cells were dissociated with Versene (Thermo Fisher Scientific #15040066) 24 h post transfection. Macroscopic sodium leak (NALCN) currents were recorded at room temperature in the whole cell configuration using an Axopatch 200B amplifier (Molecular Devices). Data were acquired using the pCLAMP10 software (Molecular Devices) and an Axon Digidata 1440A digitizer (Molecular Devices) at 10 kHz. Patch pipettes were pulled from 1.5/086 borosilicate glass capillaries (Sutter Instrument CO. #BF150-86-15). Intracellular solution contained 140 mM CsCl, 10 mM CsF, 5 mM EGTA, 10 mM HEPES and 2 mM Na_2_ATP, pH was adjusted at 7.2 with CsOH, ∼ 285 mosm/L. Extracellular solution contained 150 mM NaCl, 2 mM CaCl_2_, 1.2 MgCl_2_, 10 mM HEPES, 10 mM D-(+)-glucose, pH 7.4 adjusted with NaOH, ∼ 305 mosm/L. Other extracellular solutions used in this study are (1) NMDG 150 mM, CaCl_2_ 2 mM, MgCl_2_ 1.2 mM, HEPES 10 mM, D-(+)-glucose 10 mM, the pH was adjusted at 7,4 with HCl, ∼ 315 mosm/L (2) NaCl 150 mM, CaCl_2_ 0.1 mM, MgCl_2_ 1.2 mM, HEPES 10 mM and D-(+)-glucose 10mM, pH 7,4 adjusted with NaOH, ∼ 300 mosm/L.

### Data analysis

Data were analyzed using pCLAMP10 (Molecular Devices), Excel and GraphPad Prism 10 softwares. All data are presented as the mean ± SEM, and (n) means the number of the cells. The normality was verified for each group thanks to the Shapiro-Wilk normality test. Depending on the experimental design, data were analyzed using either one- or two-ways ANOVA. The test used in each dataset analysis is specified in the figures’ legends. Post hoc analyses for multiple comparisons were performed with Benjamini, Krieger, & Yekutieli False Discovery Rate (FDR) corrections.

## RESULTS

We collected a group of 35 individuals with *de novo NALCN* variants aged from 1 month to 59 years. All individuals were born of healthy, non-consanguineous parents. The clinical data of individuals 32 and 33 were previously reported, but no electrophysiological properties were studied (26,32).

### Phenotypic description of the new reported patients

The major clinical features of our individuals are presented in **Table 1**, with a comprehensive description available in **Supplementary Table 1**.

**Table 1:** Main clinical features of all individuals. ADHD, Attention Deficit Hyperactivity Disorder; MRI, Magnetic Resonance Imaging

| Main clinical features | Individuals of the cohort |
| --- | --- |
| Facial dysmorphic features | 34/35 |
| Short neck | 14/35 |
| Contractures | 23/35 |
| Arthrogryposis multiplex congenita | 5/35 |
| Distal arthrogryposis | 7/35 |
| Cognitive delay | 28/30 (5 NA) |
| Speech delay | 31/34 (1 NA) |
| Motor delay | 32/33 (2NA) |
| Intellectual disability | 23/26 (9 NA) |
| ADHD | 14/22 (13 NA) |
| Autistic disorders | 7/22 (13 NA) |
| Seizures | 3/34 (1 NA) |
| Axial hypotonia | 20/35 |
| Ataxia | 12/30 (5 NA) |
| Spasticity | 3/32 (3 NA) |
| Abnormal movments | 8/33 (2 NA) |
| Abnormal brain MRI | 20/25 (10 NA) |
| Cardiac disorders | 4/35 |
| Respiratory disorders | 9/35 |
| Apnea | 7/35 |
| Excessive drooling | 20/35 |
| Gastroesophageal reflux disease | 13/35 |
| Swallowing difficulties | 10/35 |
| Constipation | 11/35 |
| Hernia | 3/35 |
| Feeding difficulties | 14/35 |
| Sleeping disorders | 9/35 |
| Prenatal disorders | 12/35 |

All individuals except one (individual 31) presented with facial dysmorphic features, such as hypertelorism, broad nasal bridge, anteverted nasal tip and micrognathia were variably present. Almost all the individuals presented developmental delays as cognitive delay (28/30) and/or speech delay (31/34) and/or motor delay (32/33) (**Table 1)**. They also presented with intellectual disability (ID) (22/26), excessive drooling (20/35), gastroesophageal reflux disease (13/35), feeding difficulties (14/35), constipation (11/35), inattention, autistic disorders (7/22) and attention deficit hyperactivity disorders (14/22). Contractures in large joints of the limbs occurred in 11 patients such as hip (6/11), elbows (4/11) or knees (2/11). Small and distal joint contractures presented as ulnar deviation (11/35), adducted thumbs (10/35), camptodactyly (9/35), or fingers contractures not otherwise specified (7/35). Five patients showed isolated adducted thumbs without contractures of the other fingers and hands (5/35), and eight had clubfoot (8/35). Twelve individuals did not display contractures of limbs or face, 8 presented only one area of contractures which was distal (ulnar deviation, clubfoot, isolated adducted thumbs), 6 presented distal arthrogryposis (involving fingers contractures and/or adducted thumbs and/or camptodactyly and/or ulnar deviation and/or clubfoot) and 8 presented distal and proximal contractures. Additional features included axial hypotonia (10/35), ataxia (12/35) with episodic ataxia (2/11), ataxic movements or ataxic gait (10/11) and other movement disorders (8/33) mainly dystonia or dyskinesia.

Abnormal patterns of brain MRI of the individuals 1, 6, 7, 29, 31 and 36 are presented in **Figure 1** and of the individuals 6, 17 and 29 in the **Supplementary Figures 1**. Abnormal patterns of brain MRI included mainly cerebellar atrophy, areas of leukoencephalopathy, abnormalities of corpus callosum (short, thin and/or dysmorphic) and brain atrophy.

**Figure 1:**
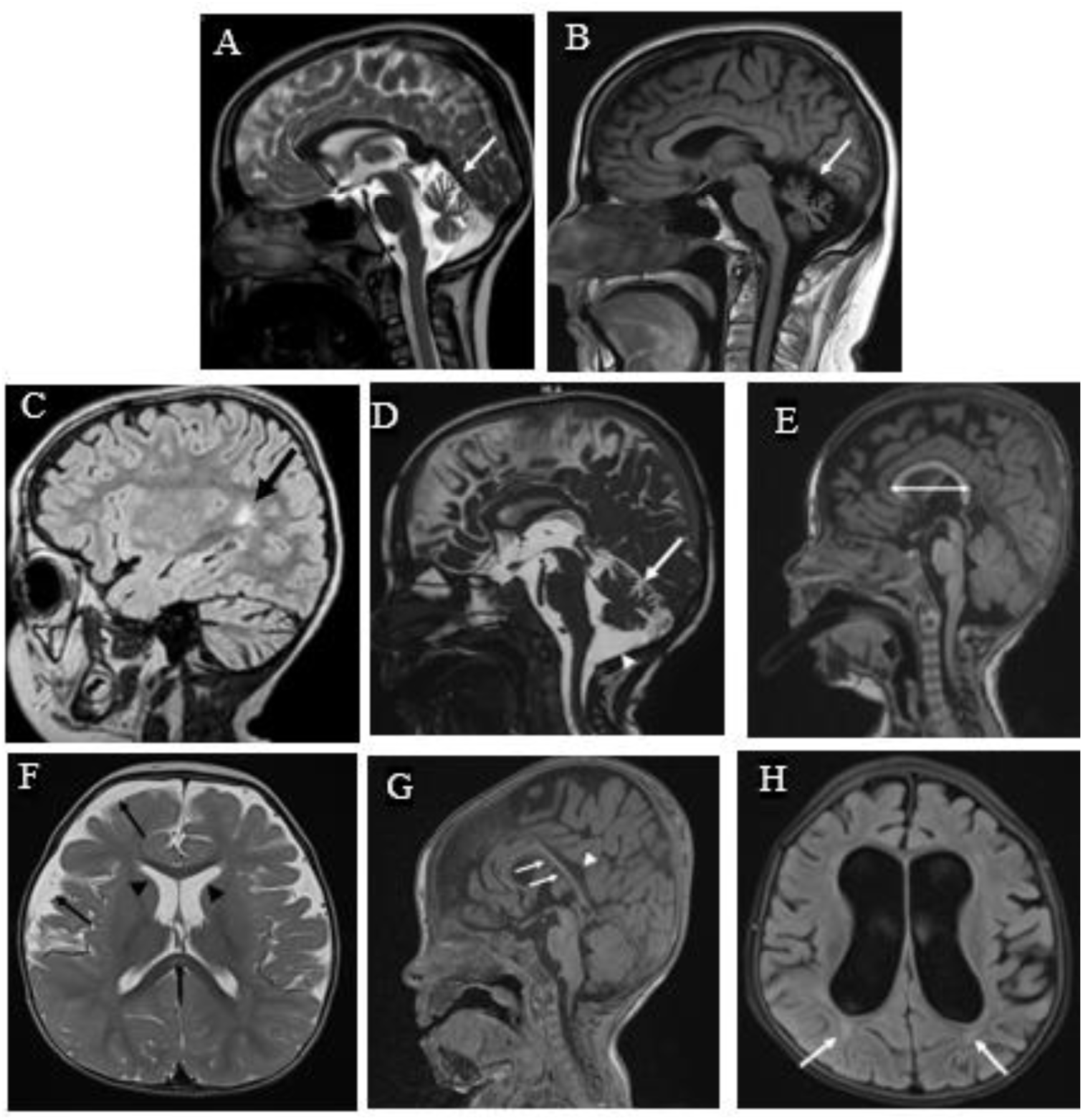
Abnormal patterns of brain MRIs of some individuals with *de novo NALCN* variants. 1 and 2 (respectively individuals 1 and 6): atrophy of both cerebellar hemispheres (white arrows) 3 (individual 27): Areas of leukoencephalopathy with FLAIR hypersignal within the periventricular regions and semi-oval centers (black arrow). 4 (individual 27): In the supratentorial area: cerebral atrophy and in the posterior cranial fossa: widening of the cerebello-medullary cistern (white arrowhead), vermian atrophy, and atrophy of both cerebellar hemispheres (white arrows). 5 and 6 (individual 29): short corpus callosum. Widening of the pericerebral spaces and frontal horns of the lateral ventricles (black arrowheads), indicative of cortico-subcortical atrophy predominantly in the fronto-parieto-temporal regions (black arrows) 7 (individual 34): Dysgenetic corpus callosum (shows a short anteroposterior diameter with overall thinning). The body, isthmus, and splenium are thin and dysmorphic (white arrows). Widening of the corpus callosum sulcus (white arrowhead) 8 (individual 34): Diffuse cortico-subcortical atrophy. Dilation of the ventricular system and cortical sulci, reflecting a decrease in white matter volume. Deep periventricular leukoencephalopathy and radial crowns with FLAIR hypersignal (white arrows).

### NALCN molecular analysis of the newly reported individuals

Of a total of 35 patients, 34 had missense variants and 1 had an in-frame insertion/deletion c.1527_1528delinsTT - p.L509_V510delinsFF. The c. 3553G>A – p.A1185T variant was identified in three individuals, and the c.986G>C – p.R329T, c.3970C>T – p.L1324F and c.3542G>A – p.R1181Q variants were identified in two individuals. The variant c.986G>C – p.R329T was identified in twins (individuals 26 and 27). Among the 28 different pathogenic *de novo* missense variants, five were already listed in the literature (*i.e.*, p.R329T, p.V316M, p.I1434V, p.L1324F, p.A1185T and p.R1181Q) (**Supplementary Table 1)**. We conducted electrophysiological recordings of 15 variants of them. Selected variants were mainly localized in the segment 6 (S6) of domain I, in S4, S5 and S6 of domains II and IV and in the P loops of domains I and III, I-II loop and III-IV loop of the NALCN protein. Mapping of the *NALCN* variants studied is presented in **Figure 2A**.

**Figure 2:**
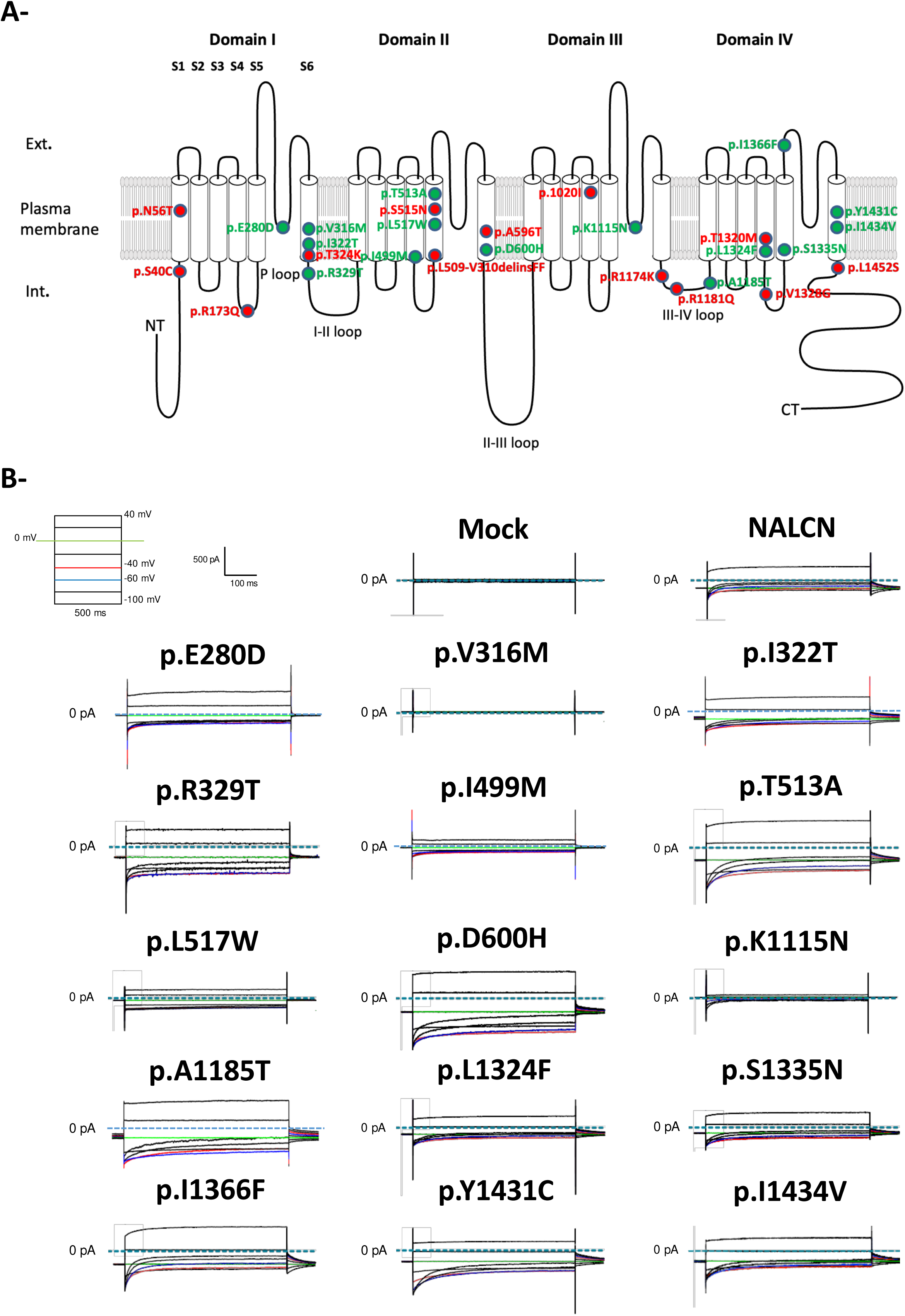
Functional expression of the NALCN channelosome in HEK-293T cells. Illustration of the general locations of 28 *de novo* pathogenic variants of *NALCN* that were investigated in this study. Variants reproduced for *in vitro* functional studies are highlighted in green. (A) Representative current traces elicited by a membrane voltage-step protocol (tests pulses from −100 mV to +40 mV) from a HP at 0 mV and recorded from HEK-293T cells co-transfected with either WT NALCN or 15 *de novo* variants along with its UNC-79, UNC-80 and FAM155A ancillary subunits. A Na^+^ leak current was observed with WT NALCN and all the variants with the exception of the p.V316M variant.

### Functional characterization of de novo NALCN pathogenic variants

We then investigated whether the severity of the patients’ symptoms and the spectrum variability could be partly explained by alterations of some biophysical properties of the NALCN current. To address this question, we introduced 15 pathogenic variants described above into the WT human *NALCN* cDNA for functional expression in the HEK-293T cell line (**Figure 2A**). To record NALCN current from HEK-293T cells using the patch-clamp technique in a whole cell configuration, we co-expressed *NALCN* along with its 3 ancillary subunits referred to as *UNC-79*, *UNC-80* and *FAM155A* as reported earlier (37). Representative sodium (Na^+^)-leak current recordings using a voltage-step protocol for the 15 NALCN variants, compared to mock (*i.e.,* NALCN replaced by GFP) and WT NALCN conditions, are shown in **Figure 2B**.

We could observe a NALCN current in 14 out of the 15 tested variants. Indeed, no current was detected for the p.V316M variant, similarly to the negative control condition (Mock), but the corresponding protein could be detected by both Western blotting and immunofluorescence (*data not shown*).

The current density-voltage (I/V) relationship of the steady-state NALCN current was then deduced from the voltage-step protocol shown in **Figure 2B** (**Figure 3A**). Heterologous expression of WT NALCN and its 3 ancillary subunits resulted in a voltage-sensitive current showing an inverted bell shape-like I/V relationship with an E_rev_ around +10 mV (**Figure 3A**), as previously described (33)(37). We also examined 2 key features of the NALCN current: it is mainly carried by extracellular Na^+^ at negative potentials and it is blocked by extracellular Ca^2+^ (3,4,33,37,38). Indeed, the NALCN current was significantly reduced when external Na^+^ was replaced by the large impermeant cation NMDG while it was potentiated when extracellular Ca^2+^ was decreased from 2 mM to 0.1 mM (**Figure 3A**). These data validated the use of HEK-293T cells to study the impact of *de novo* variants on the NALCN current.

**Figure 3:**
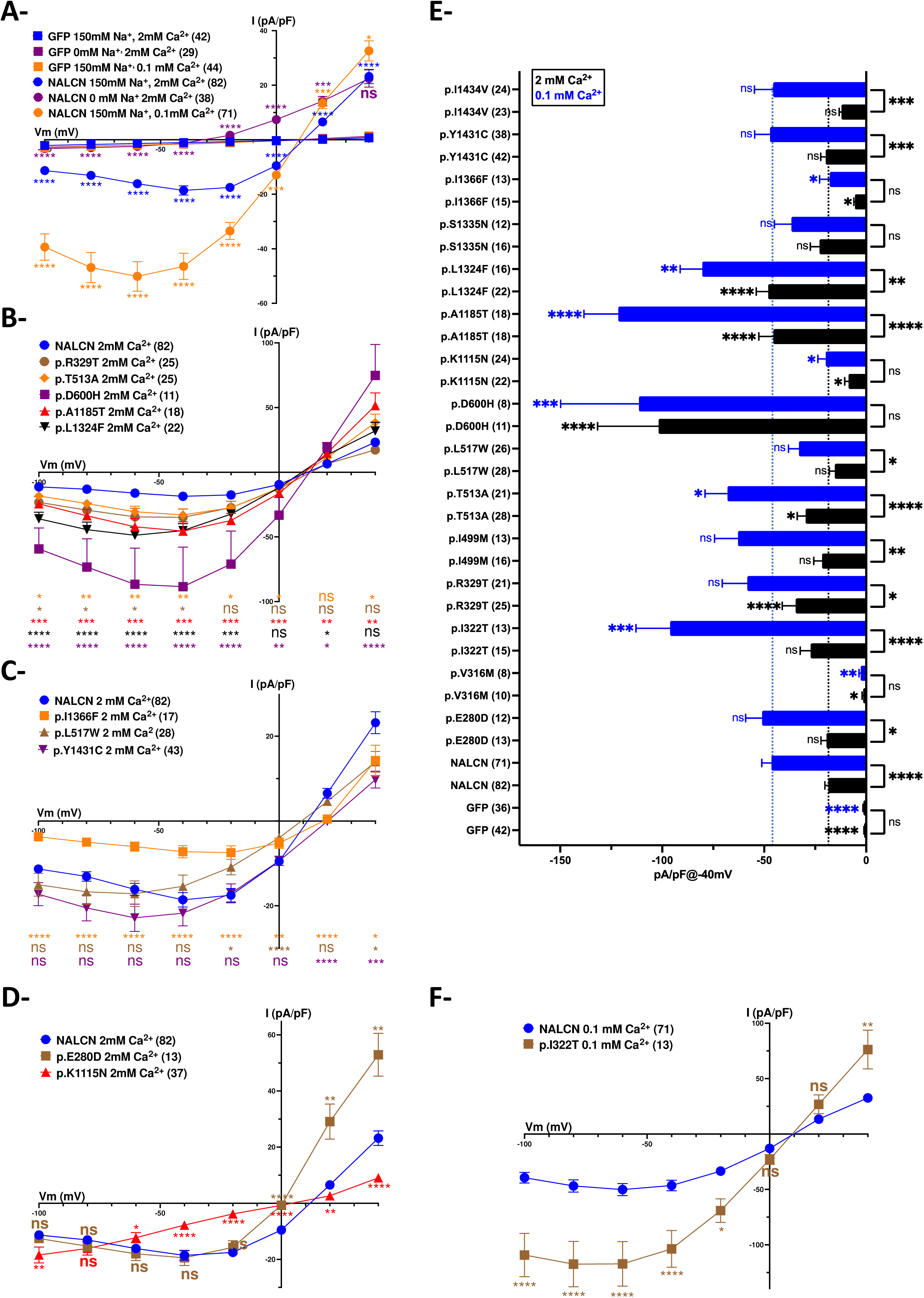
Analysis of the current densities of *de novo* pathogenic variants. (A) Intensity-Voltage relationship of the WT NALCN current in transfected HEK-293T cells. A current with an inverted bell shape-like relationship and with a reversal potential around +10 mV was observed when WT NALCN was transfected along with its UNC-79, UNC-80 and FAM155A ancillary subunits. No current was detected when NALCN was replaced by eGFP. The inward current was abolished when Na^+^ was substituted by the large impermeant cation NMDG. Conversely the current was potentiated by decreasing [Ca^2+^]_ext_ from 2 mM to 0.1 mM. (B) Intensity-Voltage relationship of gain-of-function *de novo* variants in [Ca^2+^]_ext_=2 mM. Only 5 variants out of 14 exhibited significant gain-of-function properties in terms of density of current within a negative range of membrane potential (V_m_) (*i.e.,* p.R329T, p.T513A, p.D600H, p.A1185T and p.L1324F). Most of these also showed a significant increased current density at positive potentials (*i.e.,* p.T513A, p.D600H, p.A1185T and p.L1324F). (C) Intensity-Voltage relationship of loss-of-function *de novo* variants in [Ca^2+^]_ext_=2 mM (*i.e.,* p. L517W, p.I1366F and p.Y1431C). The p.I1366F variant exhibited significant decreased current density at any tested potential while the p.L517W and p.Y1431C variants exhibited decreased current density at −20 mV/0 mV/+40 mV and +20 mV/+40 mV respectively. Note that both the p.I1366F and p.Y1431C variants showed a more depolarized E_rev_ (∼+20 mV) compared to the WT NALCN (∼+10 mV). This suggests a change in permeation properties (D) Intensity-Voltage relationship of *de novo* variants located within the selectivity filter of NALCN in [Ca^2+^]_ext_=2 mM (*i.e.,* p.E280D and p.K1115N). The p.E280D variant exhibited a significant increase in current density within positive membrane potentials (V_m_) while the p.K1115N variant exhibited a significant loss-of-function property with decreased current density for both negative and positive membrane potentials (Vm). Contrasting with WT NALCN and other *de novo* variants studied herein, the p.K1115N variant exhibited a linear Intensity-Voltage relationship. For both variants, the reversal potential E_rev_ was closed to 0 mV thus suggesting a change in permeation properties. (E) Summary of current densities of the 15 *de novo* variants compared to control conditions (*i.e.,* GFP as negative control and WT NALCN as positive control) at a membrane potential (V_m_) of −40 mV in [Ca^2+^]_ext_=2 mM and in [Ca^2+^]_ext_=0.1 mM. For [Ca^2+^]_ext_=2 mM, statistical analysis revealed a significant increase in current density for the p.R529T, p.T513A, p.D600H, p.A1185T and p.L1324F *de novo* variants while a significant decreased was observed for the p.K1115N and the p.I1366F variants as reported in panels (B), (C) and (D). For [Ca^2+^]_ext_=0.1 mM significant increase in current density was observed for the p.I322T, p.T513A, p.D600H, p.A1185T, p.L1324F variants and a decrease in current density was observed for the p.K1115N (*see* panel (D)) and the p.I1366F (*see* panel (C)) variants. Regarding the current density for the same variants in [Ca^2+^]_ext_=2 mM compared to [Ca^2+^]_ext_=0.1 mM, there was no difference for the p.D600H, p.K1115N, p.S1335N and p.I1366F variants suggesting an altered sensitivity to [Ca^2+^]_ext_. (F) Intensity-Voltage relationship of the p.I322T *de novo* variant in [Ca^2+^]_ext_=0.1 mM. The p.I322T variant exhibited significant gain-of-function property in terms of current density in [Ca^2+^]_ext_=0.1 mM but not in [Ca^2+^]_ext_=2 mM (*see* Supplementary Figure 2). The statistical significance was reached for potentials ranging from −100 mV to −20 mV as well as at +40 mV. Current amplitudes and subsequently current densities were measured at 300 msec. Statistical analysis for panels (A-D & F): two-ways ANOVA; Statistical analysis for panels (E): one-way ANOVA. Post hoc analyses for multiple comparisons were performed with Benjamini, Krieger, & Yekutieli False Discovery Rate (FDR) corrections. *P<.05, **P<.01, ***P<0.001, ****P<0.0001.

We and others have previously described gain-of-function properties of *de novo* pathogenic variants of NALCN in terms of current amplitude/density without any significant alteration in plasma membrane expression (33,34). Strikingly, only 5 out of 14 variants (*i.e.,* p.R329T, p.T513A, p.D600H, p.A1185T and p.L1324F) exhibited a significantly larger current density within negative potentials ranging from −100 to −40 mV compared to the WT NALCN (**Figure 3B**). With the exception of the p.R329T variant, the others also exhibited larger current densities at more depolarized membrane potential (Vm). These variants localize within the intracellular DI-DII linker (*i.e.,* p.R329T), pore-forming regions of domain II (*i.e.,* p.T513A, p.D600H), the intracellular DIII-DIV linker (*i.e.,* p.A1185T) and the S4 transmembrane helix of domain IV (*i.e.,* p.L1324F). Unexpectedly, we also observed a significant loss-of-function effect for the p.I1366F variants within the −100 mV to +40 mV range as well as for the p.L517W and p.Y1431C variants at more depolarized potentials (i.e., −20 mV to +40 mV) (**Figure 3C**). The reversal potential E_rev_ for p.L517W and p.Y1431C was also slightly shifted to more positive potentials compared to WT NALCN (*i.e.,* close to +20 mV compared to around +10 mV). There was no significant difference of current density at any potential for other *de novo* variants (*i.e.,* p.I322T, p.I499M, p.S1335N and p.I1434V) compared to the WT NALCN (**Supplementary Figure 2**). Intriguingly, 2 *de novo* variants with mutations in the selectivity filter of the pore-forming regions of Domain I (*i.e.,* p.E280D) and Domain III (*i.e.,* p.K1115N) were identified. This results in a selectivity filter originally composed of p.E280-p.E554-p.K1115-p.E1389 to p.D280-p.E554-p.K1115-p.E1389 and p.E280-p.E554-p.N1115-p.E1389 respectively. Such changes are predicted to putatively affect ion permeability/selectivity properties. Indeed, the p.E280D variant exhibited significantly larger current density for potentials ranging from 0 mV to +40 mV while the p.K1115N variant exhibited a loss-of-function property both at negative and positive potentials ranging from −100 mV to +40 mV (**Figure 3D**). Moreover, the I/V relationship for this latter variant was the best described with a linear shape (Slope=0.193, R squared > 0.99) suggesting a key role of the p.K1115 amino acid in providing voltage-dependence ability in recombinant system. In both cases, the reversal potential E_rev_ was slightly shifted to more negative potentials compared to WT NALCN (*i.e.,* close to 0 mV compared to around +10 mV).

It was previously described that the NALCN current is blocked by extracellular calcium (Ca^2+^) in different cell types (3,4,33,37,38) (**Figure 3A**). Therefore, we explored whether this property was conserved in our set of *de novo* variants (**Figure 3E**). With the exception of p.D600H, p.K1115N, p.S1335N and p.I1366F, we found that each tested variant was significantly sensitive to extracellular calcium at −40 mV as revealed by comparing the current densities recorded in [Ca^2+^]_ext_=2 mM and in [Ca^2+^]_ext_=0.1 mM (**Figure 3E**). Notably, we also observed that the p.I322T variant exhibited a significantly increased current only in low extracellular calcium concentration at potentials ranging to 100 mV to −20 mV and at +40 mV (**Figure 3E, 3F**). This latter finding suggested that for this particular variant, a defect in the regulation of NALCN by extracellular calcium concentration could induce a pathological state in humans. For the other variants, when compared to the WT NALCN in low extracellular calcium, 7 variants exhibited gain-of-function properties at various potentials (*i.e.,* p.E280D, p.T513A, p.D600H, p.A1185T, p.L1324F, p.Y1431C, and p.I1434V), 3 exhibited loss-of-function properties (*i.e.,* p.L517W, p.K1115N and p.I1366F) and 3 did not display any significant difference (p.R329T, p.I499M and p.S1335N) (**Supplementary Figure 3**).

Similar to what was previously described in the neuronal NG108-15 cell line (33), we also observed that NALCN current elicited by a voltage-step protocol in HEK-293T cells displayed a partial time-dependent decay before reaching a steady-state (**Figure 2B**). The slower decay kinetics reported earlier for *de novo* variants, was observed only for 7 variants (*i.e.,* p.E280D, p.I322T, p.I499M, p.T513A, p.A1185T, p.Y1431C and p.I1434V; **Figure 4A, 4B**). By contrast, 5 variants exhibited faster time-dependent decay (*i.e.,* p.R329T, p.L517W, p.D600H, p.K1115N, and p.I1366F; **Figure 4B**), while 2 variants did not display any change in kinetics, compared to WT NALCN (*i.e.,* p.L1324F and p.S1335N) (**Supplementary Figure 4**). A comprehensive summary of the biophysical properties of variants on the NALCN current is shown in **Table 2**.

**Figure 4:**
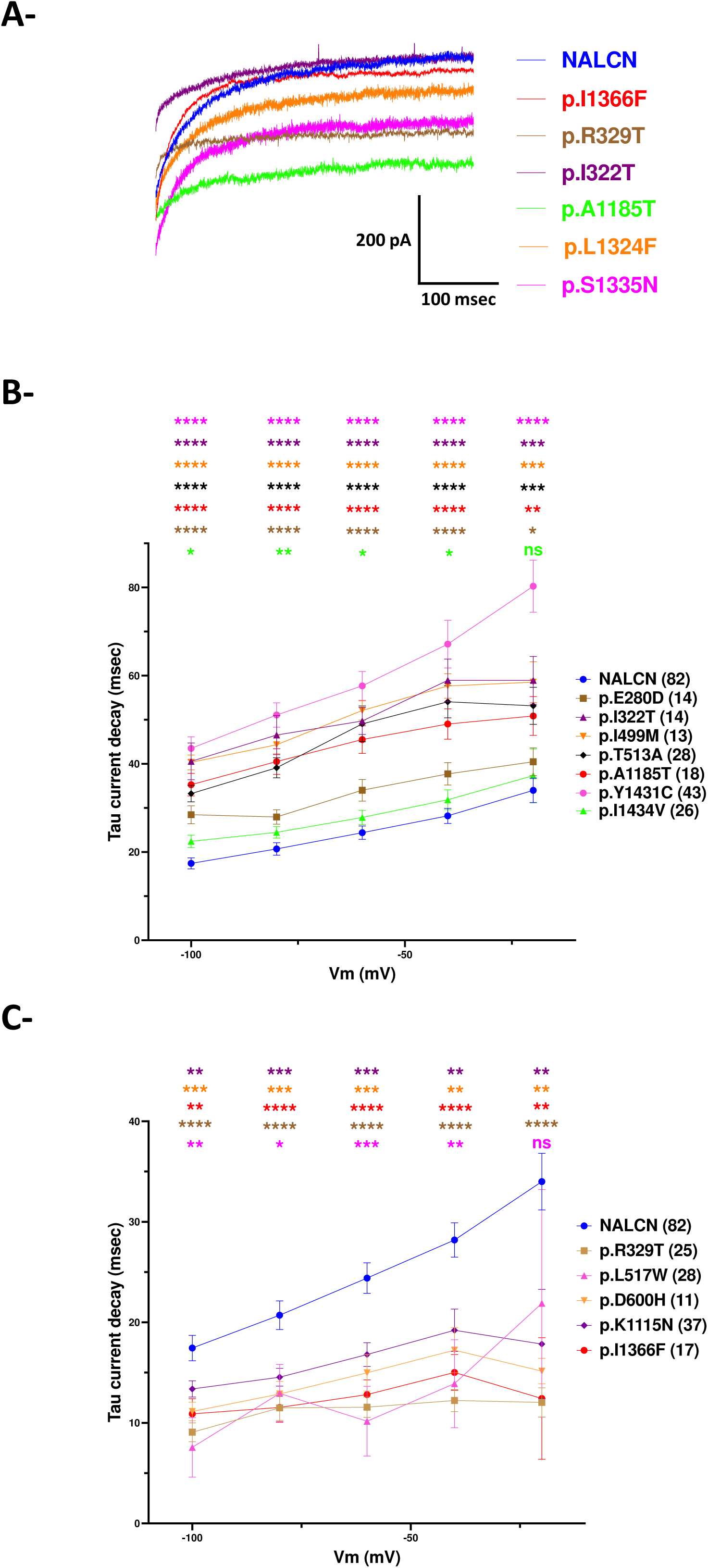
Analysis of the current decay of *de novo* pathogenic variants in [Ca^2+^]_ext_=2 mM. (A) Normalized current traces illustrating different decay kinetics of the NALCN current for the WT channel and the p.I1366F, p.R329T, p.I322T, p.A1185T, p.L1324F and p.S1335N *de novo* variants. (B) 7 out of 14 *de novo* variants significantly exhibited slower decay kinetics in [Ca^2+^]_ext_=2 mM. when compared to WT NALCN (*i.e.,* p.E280, p.I322T, p.I499M, p.T513A, p.A1185T, p.Y1431C, p.I1434V). With the exception of the p.I1434V variant, the statistical significance was reached for all tested potentials. (C) 5 out of 14 *de novo* variants significantly exhibited faster decay kinetics kinetics in [Ca^2+^]_ext_=2 mM when compared to WT NALCN (*i.e.,* p.R329T, p.L517W, p.D600H, p.K1115N, p.I1366F). With the exception of the p.L517W variant, the significance was not reached at any tested potentials. Statistical analysis for panels (B) and (C): two-ways ANOVA. Post hoc analyses for multiple comparisons were performed with Benjamini, Krieger, & Yekutieli False Discovery Rate (FDR) corrections. *P<.05, **P<.01, ***P<0.001, ****P<0.0001.

**Table 2:**
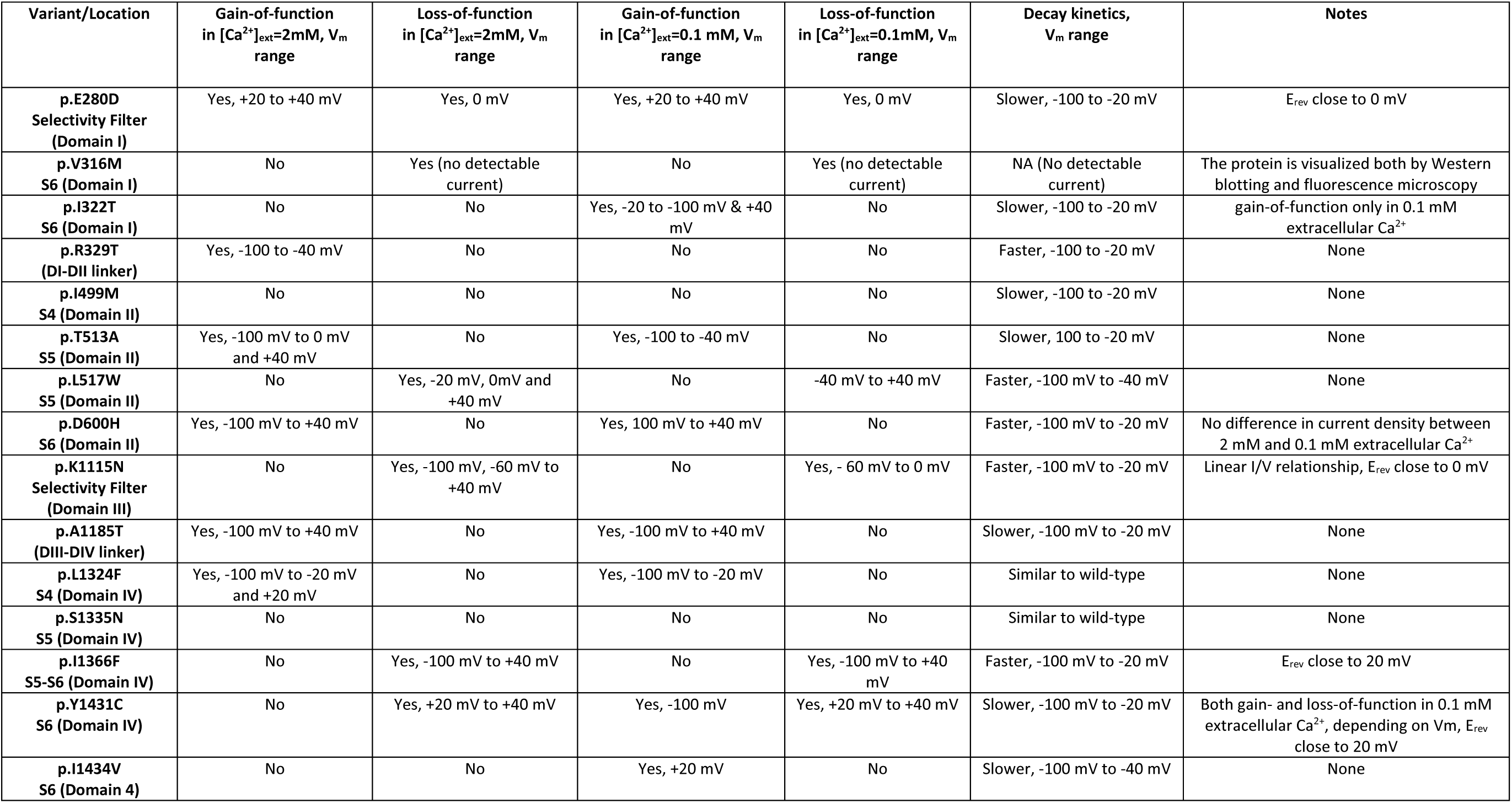
Summary of observed impact of *de novo* variants on NALCN current.

## DISCUSSION

Here, we report a large variability in the clinical spectrum in individuals with *de novo NALCN* variants, extending the original CLIFAHDD presentation. Chong et *al*, 2015 reported several *de novo NALCN* variants as causative for CLIFAHDD syndrome (39). In Chong et *al*., the continued existence of severe contractures and arthrogryposis is explained by the fact that the population analyzed by exome sequencing, revealing pathogenic de novo *NALCN* variants; was composed of individuals specifically exhibiting arthrogryposis multiplex congenita (39). The pathogenic variants reported by Chong et *al*, were all missense variants and predicted to mainly alter the S5 and S6 pore-forming segments of transmembrane domains I, II and III of *NALCN* as well as the intracellular loop connecting transmembrane domains III and IV. The more recently published de *novo NALCN* cases also exhibited limb contractures or various forms of arthrogryposis (17,22–25,27–31,40,41) with the exception of 2 cases (42,43). In contrast with previous publications, we report a large proportion of individuals with no contractures (12/35). In our study, the predominant clinical features were developmental delays, associated in 20/25 individuals with brain morphological abnormalities as cerebellar atrophy, which is also the most common pathological morphological pattern in the *de novo NALCN* variants in the literature. Axial hypotonia and ataxia appears to occur at a much higher frequency than previously described. Cerebellar ataxia was presented in the individuals with *NALCN* variants located in the selectivity filter of domain I (p.E280D), in domain II (p.T513A), in the intracellular loop linking transmembrane domains III and IV (p.A1185T), in the domain III (p.V1020I) and in the domain IV (p.S1335N, and p.Y1431C). Although described only once in the literature in the individual with the p.I1017S localized within the segment S3 of domain III of NALCN (22), dyskinesia or dystonia were observed here in 4 individuals (**Supplementary Table 1**). Moreover, areas of leukoencephalopathy were identified in 2 individuals, abnormalities of corpus callosum (as dysmorphic, thin and/or short) in 3 individuals MRI pattern which were not yet described in the literature **(Figure 1 and Supplementary Figures 1**). Overall, neurological symptoms and developmental delay appear to be the most frequent symptoms caused by *de novo NALCN* variants, whereas contractures and other limbs abnormalities are not necessarily present and may not be pathognomonic in CLIFAHDD syndrome.

Our study also points to the large heterogeneity of biophysical alterations observed with *de novo* variants, compared to the WT NALCN channel. We reported earlier that the *de novo* variants p.Y578S and p.L509S exhibited higher current densities and slower time-dependent decay kinetics, compared to WT NALCN current, independently of any impact on cell surface expression thus supporting gain of channel activity when expressed in the neuronal NG108-15 cell line (33). This gain-of-function property was then confirmed for a larger panel of *de novo* variants expressed in *Xenopus laevis* oocytes (34). However, contrasting with these previous reports, only a few *NALCN* variants out of the 15 *de novo* variants evaluated here did exhibit such gain-of-channel activity. Increase in current density in a negative range of Vm was observed for only for 5 *de novo* variants (*i.e.,* p.R329T, p.T513A, p.D600H, p.A1185T and p.L1324F). The p.R329T variant exhibited gain-of-function properties only in regular extracellular Ca^2+^ but did not display any significant difference with the WT NALCN channel in low extracellular Ca^2+^. In addition, 1 variant exhibited an increased current density only when the block by extracellular calcium was relieved (*i.e.,* p.I322T). This raises the possibility that a part of the pathogenicity of this latter variant may involve a dysregulation of the NALCN function by extracellular calcium. This variant also exhibited slower time-dependent current decay, potentially contributing to the gain-of-function property as we previously described for the p.Y578S or p.L509S *de novo* variants (33). However, we observed a diversity of changes in this time-dependent current decay. Indeed, only 7 variants showed a slower decay kinetics (*i.e.,* p.E280D, p.I322T, p.I499M, p.T513A, p.A1185T, p.Y1431C and p.I1434V) while 5 variants exhibited faster decay kinetics (*i.e.,* p.R329T, p.L517W, p.D600H, p.K1115N, p.I1366F and p.I1434V) and 2 variants did not display any alteration compared to WT NALCN (*i.e.,* p.L1324F and p.S1335N). Overall, these findings identify that the *de novo* variants exhibit contrasting electrophysiological behaviors, distinct from the originally described gain of channel activity exemplified by the p.Y578S and p.L509S *de novo* variants.

Surprisingly, we observed electrophysiological properties for a few *de novo* variants that would support a loss-of-function property, *i.e.,* the lower current density as for the p.K1115N variant. A loss-of-function property was also observed for the p.I1366F, p.L517W and p.Y1431C variants. A drastic loss of channel activity was observed with the p.V316M *de novo* variant as no NALCN current could be recorded although the corresponding protein was detectable. It remains to be determined how this variant specifically contributes to a disease state in humans.

To date, it is described that individuals with mono-allelic inherited loss-of-function variants of *NALCN* do not exhibit any obvious symptoms although this notion was recently challenged (*reviewed in* (6) *and see* (21)). In this context, it seems unlikely that the p.V316M variant exerts its pathogenic effect by just being non-conducting. We previously demonstrated that mutants and misfolded forms of the Ca_v_2.1 channels, another member of the 4-domain ion channel family, could interact with the full-length channel and induce its degradation by the proteasome (44). Such a mechanism could occur with the p.V316M variant on *NALCN*. In keeping with this idea, we previously provided some evidence this process could occur for *NALCN* when the WT channel is co-expressed with *de novo* variants in HEK-293T cells (39). However, *UNC-79*, *UNC-80* and *FAM155A* were not co-transfected in these experiments. The same hypothesis could be extendable to the other loss-of-function variants we identified either in regular extracellular calcium and/or in low extracellular calcium (*i.e.,* p.L517W, p.K1115N, p.I1366F, p.Y1431C) as well as for the p.S1335N variant that was similar to the WT channels with respect to the examined parameters. However, regarding the p.K1115N variant, the lack of voltage-dependency and changes in its permeation properties could also contribute to the development of a disease state in the corresponding patient. Further studies should investigate how loss-of-function properties of some *de novo* variants induce a pathological state. In any case, further investigations will help to decipher mechanism(s) involved in the pathogenicity of loss-of-function *de novo* variants.

In this study, we provide evidence for a large spectrum of symptoms with variable severity in patients carrying *de novo NALCN* variants, as well as a diversity of functional consequences of these variants on the NALCN current. This raise the question whether a correlation genotype-phenotype could be achieved in these patients. The variants p.Y578S and p.L509S associated with the classical CLIFAHDD phenotype (33) exhibited NALCN current with high current density and slow time-dependent decay kinetics, supporting gain-of-function behavior. Here, the variants p.R329T, p.T513A, p.D600H, p.A1185T and p.L1324F showing increased current density would add to this variants’ group. These novel variants, except p.A1185T, caused clinical features similar to the standard CLIFAHDD syndrome with arthrogryposis and developmental delays at the first plan. One of these variants (p.L1324F) associated with a more severe phenotype (individual 5, suffering from congenital arthrogryposis multiplex, and individual 32, who died *in utero* during the second trimester), while another variant (p.A1185T) was found in 2 individuals with milder phenotypes, including individual 16 without any contracture of limbs and face and no cognitive delay. Moreover, several other *de novo* variants tested here did not exhibit gain-of-function ones in regular conditions (*i.e.,* p.E280D, p.I322T, p.I499M, p.S1335N, p.Y1431C and p.I1434V). The individuals with the variants p.E280D, p.I322T, p.I499M and p.I1434V presented limbs abnormalities less severe than the classical CLIFAHDD syndrome, with for example only distal contractures. The individuals with the variants p.S1335N and p.Y1431C presented no contractures of limbs and face but a predominant phenotype of developmental delay and episodic ataxia. In our cohort, the 2 patients with the p.V316M variant (with no NALCN current detected) exhibited developmental delays, mild dysmorphic face features, respiratory and digestive disorders but no contractures for one and isolated clubfoot for the other. Three variants were considered as loss-of-function in regular extracellular calcium (*i.e.*, p.K1115N, p.L517W and p.I1366F). The 3 individuals presented similar phenotypes with predominant phenotype of developmental delays, mild facial dysmorphic features, no contractures for the individuals with variants p.K1115N and p.I1366F; but isolated clubfoot and seizures for the individual with the variant p.L517W. Our study also reveals that the individuals affected by variants exhibiting slow NALCN current decay (p.I322T, p.I499M, p.T513A, p.A1185T and p.Y1431C) presented less severe CLIFFAHD phenotypes, contrasting with that previously reported for the p.Y578S and p.L509S variants (33). In addition, the variants p.R329T and p.L1324F causing CLIFFAHD syndrome displayed respectively faster decay and no significant alteration of current decay. These results mitigate against the impact of a change in NALCN current decay in the presentation and severity of the CLIFFAHD syndrome.

### Limitations

This study documents the wide spectrum of clinical and biophysical features of *de novo NALCN* related disorders and highlights that all the *de novo* variants identified in this study caused neurodevelopmental disorders. However, a closer correlation between genotype, clinical and biophysical features could not be firmly established in this present study. Nevertheless, some patterns have emerged. The gain-of-function variants tend to cause CLIFAHDD phenotypes but with variability in the other neurological clinical features, while the variants without gain-of-function or with loss-of-function tend to cause less severe limbs abnormalities than in classical CLIFAHDD phenotype (*e.g*., with no limb and face contracture), but higher frequency of ataxia and abnormal movements. Studies of additional cases of *de novo NALCN* variants are needed to further refine the correlation between genotype, functional results and clinical phenotypes. Heterologous expression of NALCN disease variants in recipient cellular models has enabled describing their functional properties in previous studies (33,37). Here we provide electrophysiological recordings of recombinant NALCN variants using HEK-293T cells, a cellular model widely accepted for the heterologous expression of ion channels (45). Although our study shows that the properties of the NALCN current are globally well recapitulated in HEK-293T cells, the use of other cell systems could be important to model further the properties of disease variants towards refining genotype to phenotype correlations. Furthermore, the functional impact of these variants on the properties of native neurons is not yet known, and further investigations are warranted to determine the consequences of these NALCN alterations on neuronal excitability and network function. Nevertheless, this large cohort of individuals and the electrophysiological results, which have never been described in the literature, provide a better understanding about the high variability in the clinical spectrum linked to *de novo* pathogenic *NALCN* variants.

## CONCLUSION

As a conclusion, de novo *NALCN* variants invariably result in neurodevelopmental disorders. However, the associated clinical features exhibit a remarkably diverse spectrum. Importantly, not all de novo *NALCN* variants are gain-of-function, and distinct patterns in clinical phenotypes have emerged. Gain-of-function *NALCN* variants tend to cause CLIFAHDD syndrome. In contrast, non-gain-of-function or loss-of-function *NALCN* variants tend to produce with less severe disorders than the CLIFAHDD syndrome, notably lacking limb and facial contractures. The presence of other central neurological disorders is highly variable, with the possible existence of cerebellar ataxia or other movement disorders. Further studies with animal models and iPSC-derived neurons or computer modelling are needed to better understand the *in vivo* impact of these *NALCN* variants.

## ACKNOWLEDGMENTS

We express our gratitude to the non-profit patient advocacy groups that kindly helped us in distributing information about the study and that provide support to all patients and families involved, the Libellas Foundation (https://fundacionlibellas.org/) and the Channeling Hope Foundation (https://www.channelinghope.org/). This study makes use of data shared through the Gene matcher Plateform (https://genematcher.org/). We express our acknowledgements to all persons and families involved in this project as well as the Libellas foundation and the Channeling Hope Foundation. We are also very grateful to Dr HC Chua and Dr SA Pless for generously sending us the plasmids encoding the NALCN channelosome.

## CREDIT AUTHORSHIP CONTRIBUTION STATEMENT

NH: Writing – review & editing, Writing – original draft, Visualization, Validation, Supervision, Project administration, Methodology, Investigation, Formal analysis, clinical data analysis and curation, Conceptualization.

LPG and RB: Writing – review & editing, Writing – original draft, electrophysiological recordings, analysis and data curation, Investigation.

AM: Writing – review & editing, Writing – original draft, Resources, Project administration, Visualization, Validation, Supervision, Methodology, Investigation, electrophysiological analysis and data curation, Conceptualization.

CT: Writing – review & editing, Writing – original draft, Resources, Project administration, Visualization, Validation, Supervision, Methodology, Investigation and Conceptualization.

PL: Writing – review & editing, Writing – original draft, Resources, Project administration, Visualization, Validation, Supervision, Methodology, Investigation, electrophysiological analysis and data curation, Conceptualization.

PP, AGN, SW, SG, TB, MB, DN, KV, DL, KD, JD, MCN, MF, SS, EBB, JL, DY, GH, IM, CF, HW, NH, AS, LS, EA, BHL, RWG, IM, MD, GL, GR, RV, LS, AJE, FDMJ, GP, AP, SS, SRC, ER, VG, AR, MP, UH, CK, CPB, HSS, MG, FL, ES, DC, JT and LF: Contributors to the clinical data, Writing – review & editing

## DATA AVAILABILITY STATEMENT

The exhaustive data of this study are available on request to the corresponding authors.

## CODE AVAILABILITY STATEMENT

The functional data generated and analyzed during this study are within the published article and its supplementary files, and additional data are available from the corresponding author on reasonable request.

## ETHICS APPROVAL

We certify that we have received and archived written consent for participation/publication from every individual whose data is included. We certify that permissions were obtained to publish patients’ photos or MRI from all legal guardians.

