## Supplementary Figures 1-4 for "*De novo* variants of *NALCN* differentially impact both the phenotypic spectrum of patients and the biophysical properties of the NALCN current"

**Supplementary Figure 1: Abnormal patterns of brain MRIs of some individuals with *de novo NALCN* variants**

Abnormal pattern of brain MRIs of some individuals with *de novo* NALCN variants

1 to 6 respectively individual 19 (1 and 2), individual 17 (3 and 4) and individual 6 (5 and 6)

1, 3, 4, 5 et 6: Mild to moderate cerebellar atrophy

1 and 2: Mildly dilated cerebropsinal fluid spaces and brain atrophy

**Supplementary Figure 2: *De novo* pathogenic variants that do not exhibited any significant alteration in current density in $[Ca^{2+}]_{ext}$=2 mM.**

Intensity-Voltage relationship of *de novo* variants that did not displayed any significant alteration of current density with $[Ca^{2+}]_{ext}$=2 mM for potentials ranging from -100 mV to +40 mV (*i.e.,* p.I322T, p.I499M, p.S1335N and p.I1434V).

Current amplitudes and subsequently current densities were measured at 300 msec. Statistical analysis: two-ways ANOVA. Post hoc analyses for multiple comparisons were performed with Benjamini, Krieger, & Yekutieli False Discovery Rate (FDR) corrections. *P<.05, **P<.01, ***P<0.001, ****P<0.0001.

**Supplementary Figure 3: I/V relationship of *de novo* NALCN variants in $[Ca^{2+}]_{ext}$=0.1 mM.**

(A) Intensity-Voltage relationship of gain-of-function *de novo* variants in $[Ca^{2+}]_{ext}$=0.1 mM. Only 7 variants out of 14 exhibited significant gain-of-function properties in terms of current density at various range of membrane potential ($V_m$) (*i.e.,* p.E280D, p.T513A, p.D600H, p.A1185T, p.L1324F, p.Y1431C, p.I1434V).

(B) Intensity-Voltage relationship of loss-of-function *de novo* variants in $[Ca^{2+}]_{ext}$=0.1 mM. 3 variants out of 14 exhibited significant loss-of-function properties in terms of current density of at various range of membrane potential ($V_m$) (*i.e.,* p.L517W, p.K1115N, p.I1366F).

(C) Intensity-Voltage relationship of *de novo* variants that did not exhibited any significant changes in current density in $[Ca^{2+}]_{ext}$=0.1 mM. 3 out of 14 variants were identified (*i.e.,* p.R329T, p.I499M, p.S1335N).

Statistical analysis for panels (A-C): two-ways ANOVA. Post hoc analyses for multiple comparisons were performed with Benjamini, Krieger, & Yekutieli False Discovery Rate (FDR) corrections. *P<.05, **P<.01, ***P<0.001, ****P<0.0001.

**Supplementary Figure 4: D*e novo* pathogenic variants that do not exhibited any significant alteration in time-dependent decay kinetics in $[Ca^{2+}]_{ext}$=2 mM.**

2 out of 14 *de novo* variants did not display any significant decay kinetics with $[Ca^{2+}]_{ext}$=2 mM when compared to WT NALCN (*i.e.,* p.L1324F, p.S1335N).

Statistical analysis: two-ways ANOVA. Post hoc analyses for multiple comparisons were performed with Benjamini, Krieger, & Yekutieli False Discovery Rate (FDR) corrections. *P<.05, **P<.01, ***P<0.001, ****P<0.0001.

**Supplementary Table 1**: **Exhaustive presentation of the clinical features of all individuals**

ADHD, Attention Deficit Hyperactivity Disorder

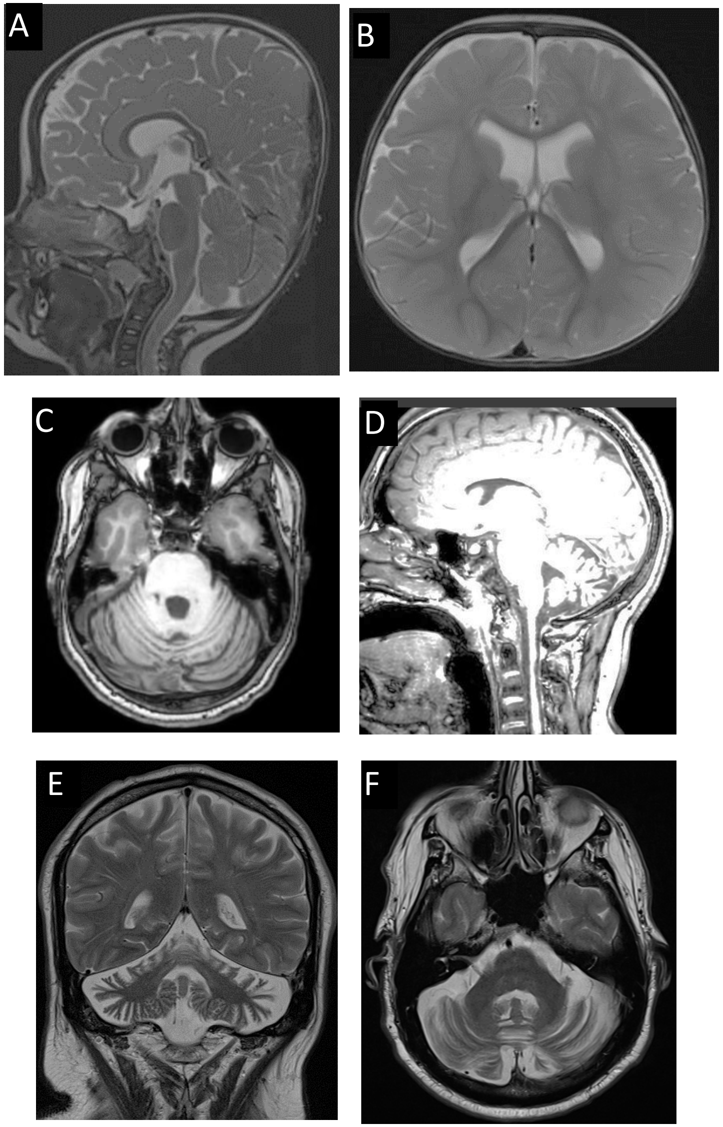


**Supplementary Figure 1**

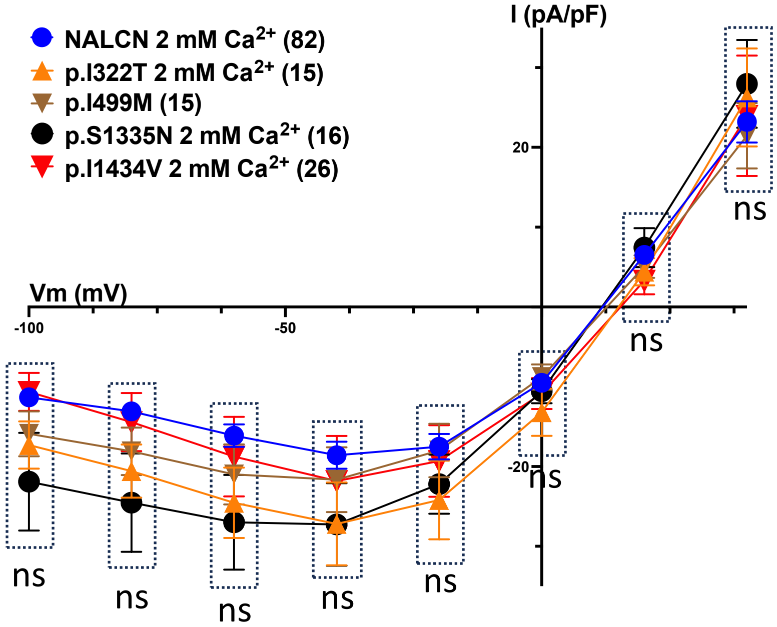


**Supplementary Figure 2**

A-

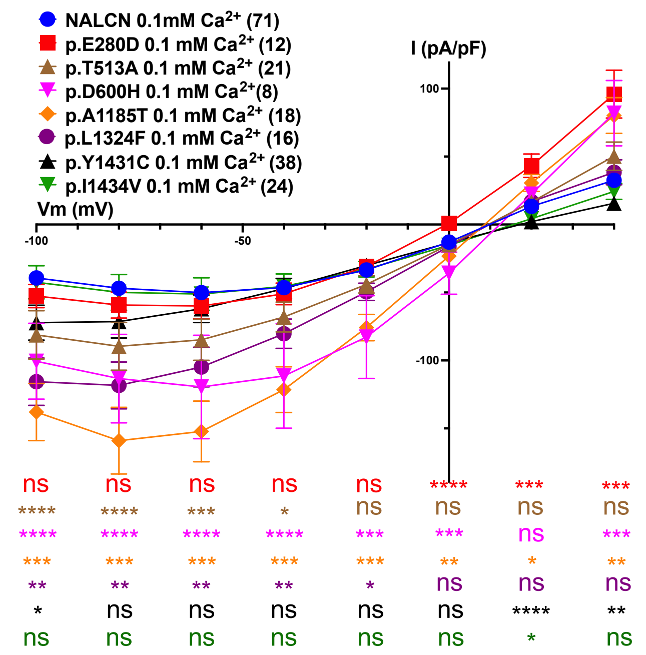


B-

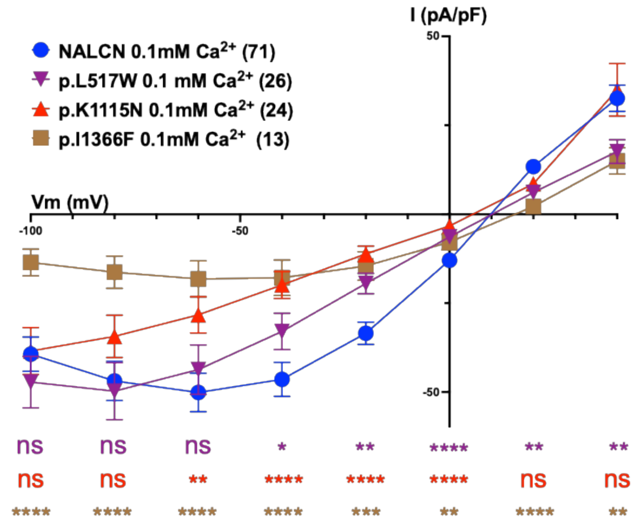


C-

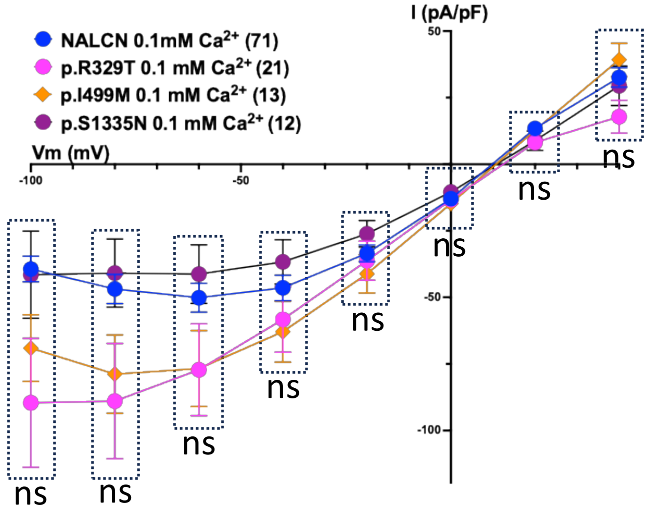


Supplementary Figure 3

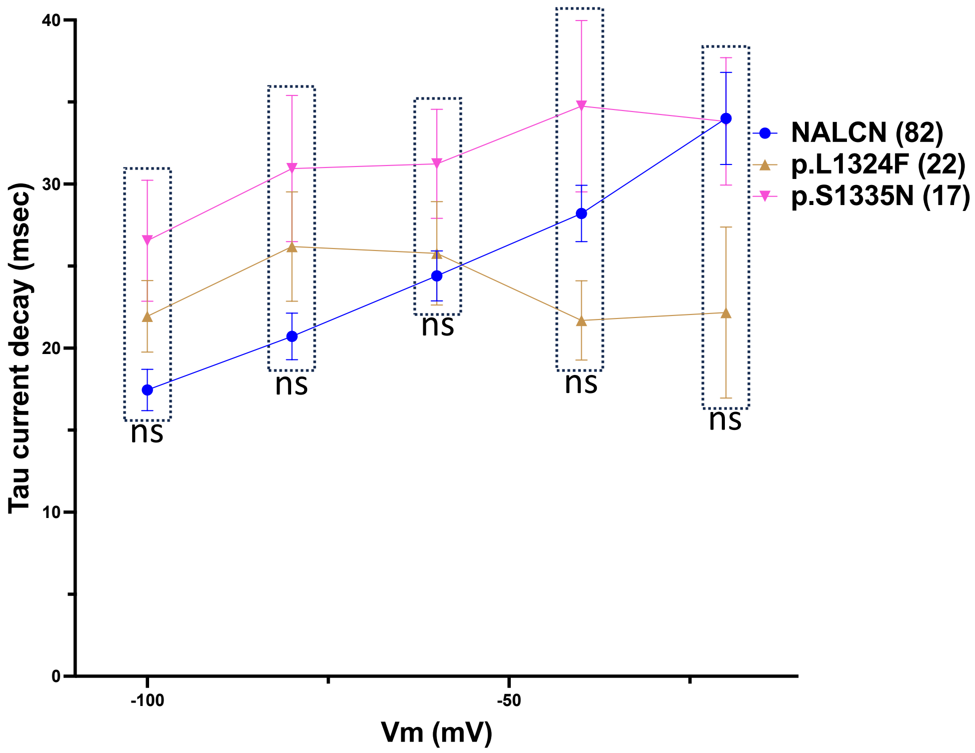


**Supplementary Figure 4**
